# Stay-at-home policy: is it a case of exception fallacy? An internet-based ecological study

**DOI:** 10.1101/2020.10.13.20211284

**Authors:** Ricardo F. Savaris, Guilherme Pumi, Jovani Dalzochio, Rafael Kunst

**Author notes:** Correspondence to: Serv. Ginecologia e Obstetrícia, Hospital de Porto Alegre, Rua Ramiro Barcelos 2350, Porto Alegre, RS, Brazil, CEP 90035-903. Postgraduate of BigData, Data Science and Machine Learning course, Unisinos, Porto Alegre, RS, Brasil.

## Abstract

**Background:** Countries with strict lockdown had a spike on the number of deaths. A recent mathematical model has suggested that staying at home did not play a dominant role in reducing COVID-19 transmission. Comparison between number of deaths and social mobility is difficult due to the non-stationary nature of the COVID-19 data.

**Objective:** To propose a novel approach to assess the association between staying at home values and the reduction/increase in the number of deaths due to COVID-19 in several regions around the world.

**Methods:** In this ecological study, data from www.google.com/covid19/mobility/, ourworldindata.org and covid.saude.gov.br were combined. Countries with >100 deaths and with a Healthcare Access and Quality Index of ≥67 were included. Data were preprocessed and analyzed using the difference between number of deaths/million between 2 regions and the difference between the percentage of staying at home. Analysis was performed using linear regression and residual analysis

**Results:** After preprocessing the data, 87 regions around the world were included, yielding 3,741 pairwise comparisons for linear regression analysis. Only 63 (1.6%) comparisons were significant.

**Discussion:** With our results, we were not able to explain if COVID-19 mortality is reduced by staying as home in ∼98% of the comparisons after epidemiological weeks 9 to 34.

## Introduction

By late September, 2020, approximately one million people worldwide had died from the new coronavirus (COVID-19) (Coronavirus Update (Live): 13,578,330 Cases and 583,696 Deaths from COVID-19 Virus Pandemic - Worldometer). Wearing masks, taking personal precautions, testing for COVID-19 and social distancing have been advocated for controlling the pandemic (Huang and Chen 2020; Lin et al. 2020; Wu and Qi 2020). To achieve source control and stop transmission, social distancing has been interpreted by many as staying at home. Such policies across multiple jurisdictions were suggested by some experts (Guest et al. 2020). These measures were supported by the World Health Organization (WHO Director-General’s opening remarks at the media briefing on COVID-19 - 13 April 2020), local authorities (Ministry of Housing, Communities and Local Government 2020; Mucientes and Carrasco 2020; Governor Cuomo Signs the “New York State on PAUSE” Executive Order 2020), and encouraged on social media platforms (Criativo; A Movement to Stop the COVID-19 Pandemic | #StayTheFuckHome, #[stayathome] (Brazilian twitter)).

Some mathematical models and meta-analyses have shown a marked reduction in COVID-19 cases (Ambikapathy and Krishnamurthy 2020; Espinoza et al. 2020; Ibarra-Vega 2020; Liu et al. 2020; Nussbaumer-Streit et al. 2020; Sjödin et al. 2020) and deaths (Ferguson et al. 2020; Semenova et al. 2020) associated with lockdown policies. Brazilian researchers have published mathematical models of spreading patterns (Peixoto et al. 2020) and suggested implementing social distancing measures and protection policies to control virus transmission (Aquino et al. 2020). By May 5th, 2020, an early report, using number of curfew days in 49 countries, found evidence that lockdown could be used to suppress the spread of COVID-19 (Atalan 2020). Measures to address the COVID-19 pandemic with Non-Pharmacological Interventions (NPIs) were adopted after Brazil enacted Law No. 13979 (Imprensa Nacional), and this was followed by many states such as Rio de Janeiro (Decreto 46970 27/03/2020), the Federal District of Brasília (Decree No. 40520, dated March 14^th^, 2020) (Decreto 40520 de 14/03/2020), the city of São Paulo (Decree No. 59.283, dated March 16^th^, 2020) (Decreto 59283 2020 de São Paulo SP), and the State of Rio Grande do Sul (Decree No. 55240/2020, dated May 10^th^, 2020) (Decreto 55240 de 10/05/2020). It was expected that, with these actions, the number of deaths by COVID-19 would be reduced. Of note, the country’s most populous state, São Paulo, adopted rigorous quarantine measures and put them into effect on March 24^th^, 2020 (Decreto 59283 2020 de São Paulo SP). Internationally, Peru adopted the world’s strictest lockdown (Tegel 2020).

Recently, Google LLC published datasets indicating changes in mobility (compared to an average baseline before the COVID-19 pandemic). These reports were created with aggregated, anonymized sets of daily and dynamic data at country and sub-regional levels drawn from users who had enabled the Location History setting on their cell phones. These data reflect real-world changes in social behavior and provide information on mobility trends for places like grocery stores, pharmacies, parks, public transit stations, retail and recreation locations, residences, and workplaces, when compared to the baseline period prior to the pandemic (Google LLC). Mobility in places of residence provides information about the “time spent in residences”, which we will hereafter call “staying at home” and use as a surrogate for measuring adherence to stay-at-home policies.

Studies using Google COVID-19 Community Mobility Reports and the daily number of new COVID-19 cases have shown that over 7 weeks a strong correlation between staying at home and the reduction of COVID-19 cases in 20 counties in the United States (Badr et al. 2020); COVID-19 cases decreased by 49% after 2 weeks of staying at home (Banerjee and Nayak 2020); the incidence of new cases/100,000 people was also reduced (Wang et al. 2020); social distancing policies were associated with reduction in COVID-19 spread in the US (Gao et al. 2020); as well as in 49 countries around the world (Atalan 2020). A recent report using Brazilian and European data has shown a correlation between NPI stringency and the spread of COVID-19 (Candido et al. 2020; Islam et al. 2020); these analyses are debatable, however, due to their short time span and the type of time series behavior (Bernal et al. 2017), or for their use of Pearson’s correlation in the context of non-stationary time series (Gao et al. 2020). For instance, applying the same statistical analysis to stationary and non-stationary time series is not sufficient for statistical analysis (Nason 2006), and the latter is the case with this COVID-19 data. A 2020 Cochrane systematic review of this topic reported that they were not completely certain about this evidence for several reasons. The COVID-19 studies based their models on limited data and made different assumptions about the virus (Nussbaumer-Streit et al. 2020); the stay-at-home variable was analyzed as a binary indicator (Sen et al. 2020); and the number of new cases could have been substantially undocumented (Li et al. 2020); all which may have biased the results. A sophisticated mathematical model based on a high-dimensional system of partial differential equations to represent disease spread has been proposed (Zamir et al. 2020). According to this model, staying at home did not play a dominant role in disease transmission, but the combination of these, together with the use of face masks, hand washing, early-case detection (PCR test), and the use of hand sanitizers for at least 50 days could have reduced the number of new cases. Finally, after 2 months, the simulations that drove the world to lockdown have been questioned (Boretti 2020).

After more than 25 epidemiological weeks of this pandemic, verifying if staying at home had an impact on mortality rates is of particular interest. A PUBMED search with the terms “COVID-19” AND (Mobility) (search made on September 8th, 2020) yielded 246 articles; of these, 35 were relevant to mobility measures and COVID-19, but none compared mobility reduction to mortality rates.

We are looking for the association between two variables: deaths/million and the percentage of people who remained in their residences. Comparison, however, is difficult due to the non-stationary nature of the data. To overcome this problem, we proposed a novel approach to assess the association between staying at home values and the reduction/increase in the number of deaths due to COVID-19 in several regions around the world. If the variation in the difference between the number of deaths/million in two countries, say A and B, and the variation in the difference of the staying at home values between A and B present similar patterns, this is due to an association between the two variables. In contrast, if these patterns are very different, this is evidence that staying at home values and the number of deaths/million are not related (unless, of course, other unaccounted for factors are at play).

## Material and methods

### Study design

This is an ecological study using data available on the Internet.

### Setting - Data collection on mobility

Google COVID-19 Community Mobility Reports provided data on mobility from 138 countries and regions (Coronavirus Source Data 2020, Coronavírus Brasil) between February 15^th^ and August 21^st^, 2020.(Google LLC) Data regarding the average times spent at home was generated in comparison to the baseline. Baseline was considered to be the median value from between January 3^rd^ and February 6^th^, 2020. Data obtained between February 15^th^ and August 21^th^ 2020 was divided into epidemiological weeks (epi-weeks) and the mean percentage of time spent staying at home per week was obtained.

### Data collection on mortality

Numbers of daily deaths from selected regions were obtained from open databases (Coronavirus Source Data 2020, Coronavírus Brasil) on August 27^st^, 2020.

### Inclusion criteria for analysis

Only regions with mobility data and with more than 100 deaths, by August 26th, 2020, were included in this study. For data quality, only countries with Healthcare Access and Quality Index (HAQI) of ≥ 67 were included.(Barber et al. 2017) By choosing a HAQI of ≥ 67, we assumed that data from these countries were reliable and healthcare was of high quality. For Brazilian regions, a HAQI was substituted for the Human Development Index (HDI), and those with <0.549 (low) were excluded.

Three major cities with >100 deaths and well-established results (Tokyo, Japan; Berlin, Germany, and New York, USA) were selected as controlled areas.

### Dataset of COVID-19 cases and associated data

After inclusion of the countries/regions, further data were obtained to reduce comparison bias, including population density (population/km^2^), percentage of the urban population, HDI, and the total area of the region in square kilometers. All data were obtained from open databases.(2019 Human Development Index Ranking, [Cities and States Statistics], Population by Country (2020) - Worldometer)

### Classification of areas with COVID-19

Regions were classified as controlled for cases of COVID-19 if they present at least two out of the three following conditions: **a)** type of transmission classified as “clusters of cases”, **b)** a downward curve of newly reported deaths in the last seven days, and **c)** a flat curve in the cumulative total number of deaths in the last seven days (variation of 5%) according to the World Health Organization.(WHO Coronavirus Disease (COVID-19) Dashboard) An example is shown in Figure S3 (supplement).

Data from the cities (Tokyo, Berlin, New York, Fortaleza, Belo Horizonte, Manaus, Rio de Janeiro, São Paulo, and Porto Alegre) were obtained from official government sites.(Population of Tokyo - Tokyo Metropolitan Government, Berlin, COVID-19:Data, Planning-Population-Census 2010-DCP) Tokyo, Berlin and New York were chosen for having controlled the COVID-19 dissemination, for representing three different continents, and for similarity to major Brazilian cities (Fortaleza, Belo Horizonte, Manaus, Rio de Janeiro, São Paulo, and Porto Alegre).

### Merged database

Different databases from the sites mentioned above were merged using Microsoft Excel Power Query (Microsoft Office 2010 for Windows Version 14.0.7232.5000) and manually inspected for consistency.

### Processing the data - cleaning

Data collected from multiple regions were processed using Python 3.7.3 in the Jupyter Notebook environment through the use of the Python Data Analysis Library in Google Colab Research. Details of preprocessing are described in Python script (Supplemental material). Briefly, after taking the sum of deaths/million per epi-week, and the average of the variable “staying at home” per epi-week, non-stationary patterns were mitigated by subtracting week_t_ by week_t-1_.

### Time series data setup and variables

Details regarding the pre-processing and methodological details were presented on the *approach for analyzing the time series data*. Our variables were the difference in the variation of deaths between locations A and B (dependent variable - outcome), and the difference in the variation of staying at home values between the same location (independent variable).

### Comparison between areas

Direct comparison, between regions with and without controlled COVID-19 cases, was considered in two scenarios: 1) Restrictive if, at least three out of four of the following conditions were similar: a) population density, b) percentage of the urban population, c) HDI and d) total area of the region. Similarity was considered adequate when a variation in conditions a), b), and c) was within 30%, while, for condition d), a variation of 50% was considered adequate. 2) Global: all regions and countries were compared to each other.

### Statistical analysis

#### Rationale

Time series on COVID-19 mortality (deaths/millions) display a non-stationary pattern. The daily data present a very distinct seasonal behavior on the weekends, with valleys on Saturdays and Sundays followed by peaks on Mondays (Figure S1)

To make it stationary, one may introduce dummy variables for Saturdays, Sundays, and Mondays, regress the number of deaths in these dummy variables, and then analyze the residuals. However, in most cases, the residuals are still non-stationary time series, and special treatment would be required in each case. Although this approach may be feasible for a few series, we are interested in analyzing hundreds of time series from different countries and regions. Hence, we need a more efficient way to deal with this amount of data. The covariates present another issue in regressing the daily time series of deaths/staying at home. The covariates are typically correlated with error terms due to public policies adopted by regions/countries. Mechanisms controlling social isolation are intrinsically related to the number of deaths/cases in each location. An increase in the death rate may cause more stringent policies to be adopted, which increases the percentage of people staying at home. This change causes an imbalance between the observed number of deaths and staying at home levels. In a regression model, this discrepancy is accounted for in the error term. Hence, the error term will change in accordance with staying at home levels.

#### Approach for analyzing the time series data

Data aggregation by epidemiological week is a plausible alternative (Figure S2). In this way, artificial seasonality, imposed by work scheduled during weekends and the effect of governmental control over social interaction, in a regression framework, are mitigated. The drawback is that the sample size is significantly reduced from 187 days (Figure S1) to 26 epidemiological weeks (Figure S2).

Aggregation by epidemiological week, however, still yields non-stationary time series in most cases. To overcome this problem, we differentiated each time series. Recall that if *Z*_*t*_ denotes the number of deaths in the *t*-th epidemiological week, we define the first difference of *Z*_*t*_ as

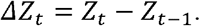

Intuitively, *ΔZ*_*t*_ denotes the variation of deaths between weeks *t* and *t-1*, also known as the flux of deaths. The same is valid for the staying at home time series. This simple operation yielded, in most cases, stationary time series, and verified with the so-called Phillips-Perron stationarity test (Perron 1988). In the few cases where the resulting time series did not reject the null hypothesis of non-stationarity (technically, the existence of a unitary root, in the time series characteristic), this was due to the presence of one or two outliers combined with the small sample size. These outliers were usually related to the very low incidence of COVID-19 deaths by the 9^th^ epidemiological week when paired with countries with a significant number of deaths in that same week, thus resulting in an outlier which cannot be accounted for by linear regression.(Perron 1988)

To investigate pairwise behavior, we propose a method to assess the relationship between deaths and staying at home data between various countries and regions. For two countries/regions, say A and B, let 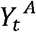 and 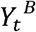 denote the number of deaths per million at epidemiological week *t* for country A and B respectively, while 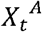 and 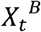 denote the staying at home at epidemiological week *t* for A and B, respectively. The idea is to regress the difference 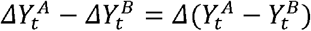 on 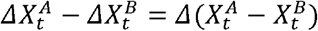. Formally, we perform the regression

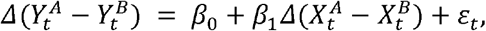

where *β*_0_ and *β*_1_ are unknown coefficients and *ε*_*t*_denotes an error term. Estimation of *β*_0_ and *β*_1_ is carried out through ordinary least squares. The interpretation of the model is important. We are regressing the difference in the variation of deaths between locations A and B into the difference in the variation of staying at home values between the same location.

If the number of deaths in locations A and B have a similar functional behavior over time, then 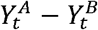 tends to be near-constant, and 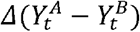 tends to oscillate around zero. If the same applies to 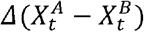, then we expect *β*_1_ ≠ 0; consequently, we conclude that the behavior, between A and B, is similar and the number of deaths and the percentage of staying at home are associated in these regions. The other non-spurious situation implying *β*_1_ ≠ 0 occurs when the variation in the number of deaths in locations A and B increases/decreases over time following a certain pattern, while the variation in the percentage of “staying at home” values also increases/decreases following the same pattern (apart from the direction). In this situation, we found different epidemiological patterns as in the variation in the number of deaths, and in the staying at home values, in locations A and B were on opposite trends. However, if these patterns were similar (proportional), this would be captured in the difference and, as a consequence, in the regression. This means that the different trends were near proportional and, hence, the variation in staying at home is associated with the variation in deaths.

The proposed approach presents a way to evaluate staying at home and the number of deaths between two countries/regions. In the section below “Definition of areas with and without controlled cases of COVID-19”, each country/region was classified into a binary class: either controlled or not controlled areas for COVID-19. The proposed method allows for insights regarding the association of the number of deaths and staying at home levels between countries/regions with similar/different degrees of COVID-19 control.

Estimation of *β*_0_ and *β*_1_ is carried out through ordinary least squares. Assumptions related to consistency, efficiency, and asymptotic normality of the ordinary least squares, in the context of time series regression, can be found in Greene, 2012 (Greene 2012). Since we are comparing many time series, to avoid any problem with spurious regression, we performed a cointegration test between the response and covariates. In this context, this is equivalent to testing the stationarity of *ε*_*t*_, which was done by performing the Phillips-Perron test. Residual analysis is of utmost importance in linear regression, especially in the context of small samples. The steps and tests performed in the residual analysis are described in the statistical analysis section.

After data preprocessing, the association between the number of deaths and staying at home was verified using a linear regression approach. Data were analyzed using the Python model statsmodels.api v0.12.0 (statsmodels.regression.linear_model.OLS; statsmodels.org), and double-checked using R version 3.6.1. False Discovery Rate proposed by Benjamini-Hochberg (FDR-BH) was used for multiple testing.

We checked the residuals for heteroskedasticity using White’s test; for the presence of autocorrelation using the Lagrange Multiplier test; for normality using the Shapiro-Wilk’s normality test; and for functional specification using the Ramsey’s RESET test. All tests were performed with a 0.05 significance level and the analysis was performed with R version 3.6.1.

Data from 30 restrictive comparisons were manually inspected and checked a third time using Microsoft Excel (Microsoft). A heat map was designed using GraphPad Prism version 8.4.3 for Mac (GraphPad Software, San Diego, California, USA). Graphs plotting the number of deaths/million and staying at home over epidemiological weeks were obtained from Google Sheets.

## Results

A flowchart of the data manipulation is depicted in Figure 1. Briefly, Google COVID-19 Community Mobility Report data between February 16^th^ and August 21^st^, 2020, yielded 138 separate countries and their regions. The website Our World in Data provided data on 212 countries (between December 31^st^, 2019, and August 26^th^, 2020), and the Brazilian Health Ministry website provided data on all states (n=27) and cities (n=5,570) in Brazil (February 25^th^ to August 26^th^, 2020).

**Figure 1.**
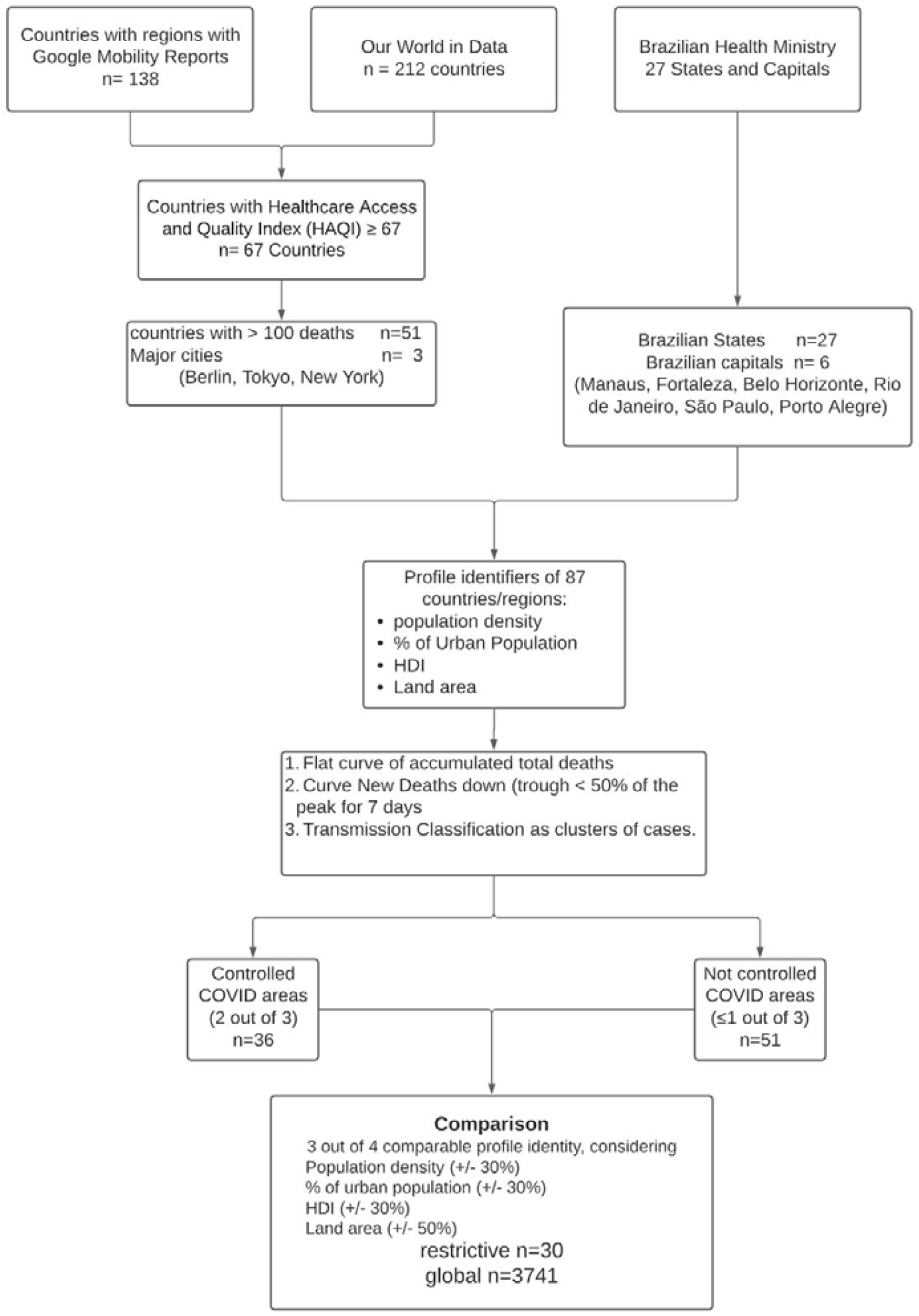
Flow chart of the data setup (Further details are in <u>Supplemental Material - Flow chart</u>).

After data compilation, a total of 87 regions and countries were selected: 51 countries, 27 States in Brazil, six major Brazilian State capitals [Manaus, Amazonas (AM), Fortaleza, Ceará (CE), Belo Horizonte, Minas Gerais (MG), Rio de Janeiro, Rio de Janeiro (RJ), São Paulo, São Paulo (SP) and Porto Alegre, Rio Grande do Sul (RS)], and three major cities throughout the world (Tokyo, Berlin and New York) (Figure 1).

Characteristics of these 87 regions are presented in Table 1 (further details are in <u>Supplemental Material - Characteristics of Regions</u>).

**Table 1.**
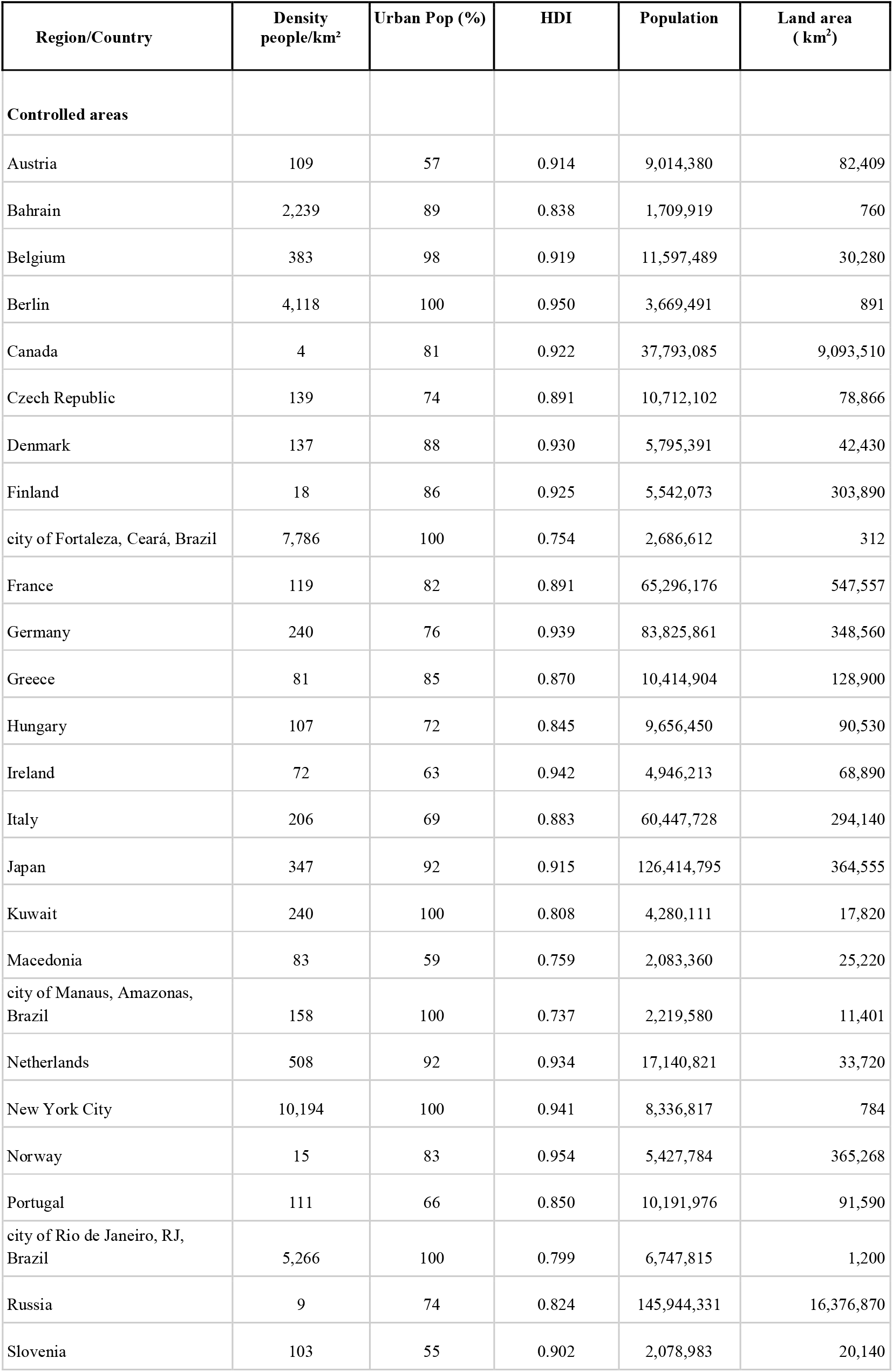
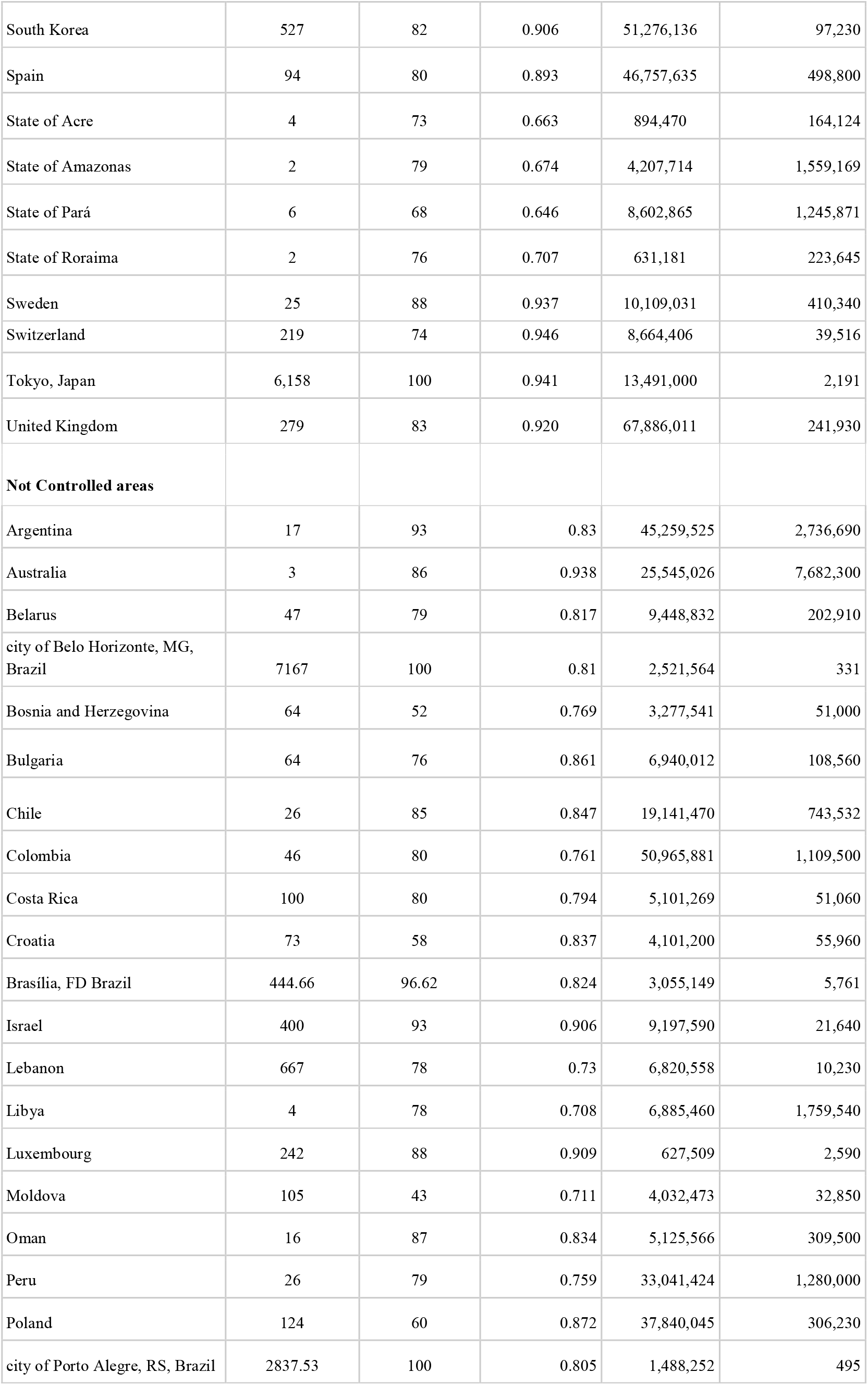
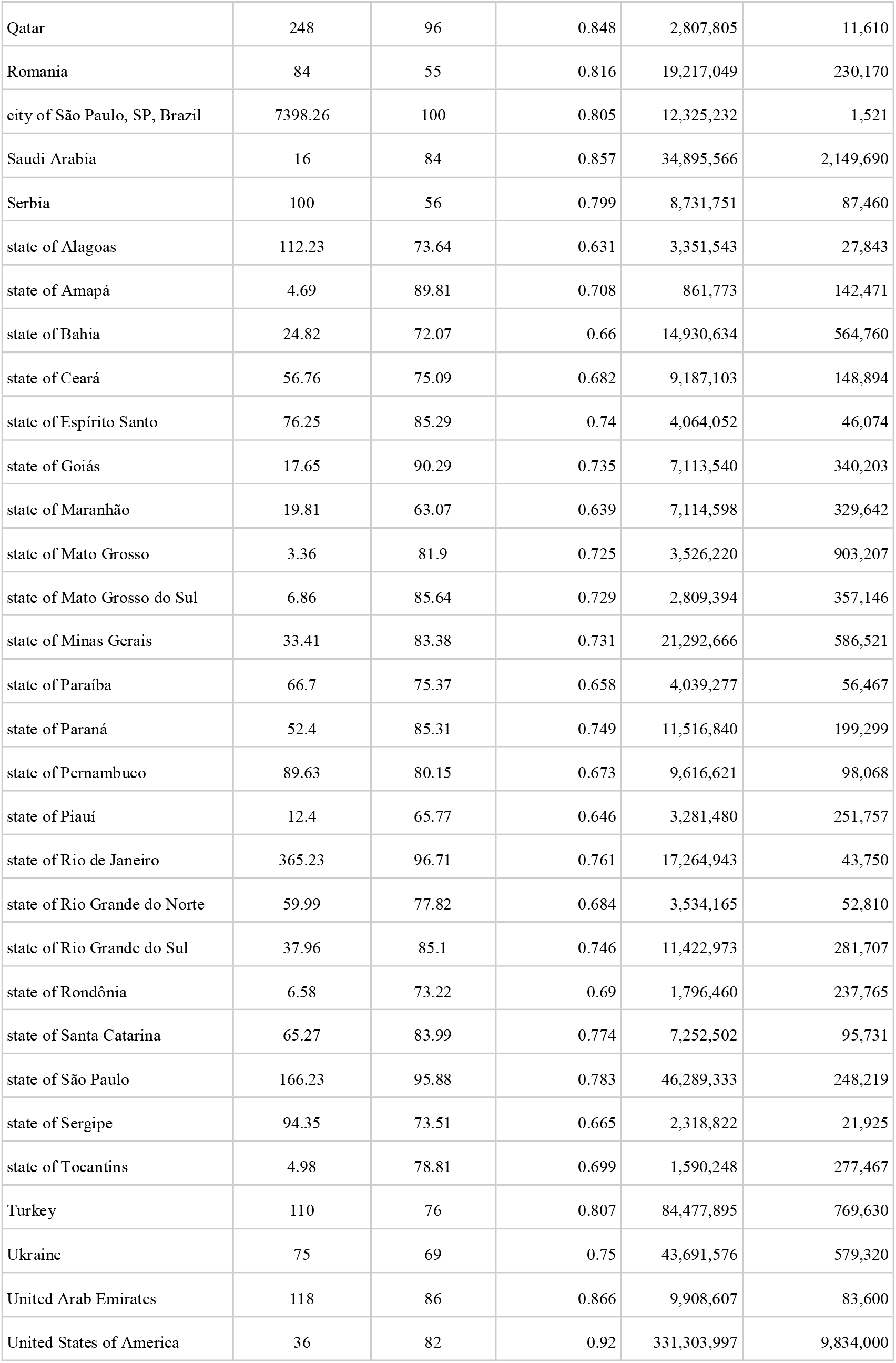
Characteristics of the 87 regions and countries used for comparison in the study. HDI = Human Development Index (the higher, the better).

| Region/Country | Density<br>people/km<sup>2</sup> | Urban Pop (%) | HDI | Population | Land area<br>( km<sup>2</sup>) |
| --- | --- | --- | --- | --- | --- |
| **Controlled areas** |  |  |  |  |  |
| Austria | 109 | 57 | 0.914 | 9,014,380 | 82,409 |
| Bahrain | 2,239 | 89 | 0.838 | 1,709,919 | 760 |
| Belgium | 383 | 98 | 0.919 | 11,597,489 | 30,280 |
| Berlin | 4,118 | 100 | 0.950 | 3,669,491 | 891 |
| Canada | 4 | 81 | 0.922 | 37,793,085 | 9,093,510 |
| Czech Republic | 139 | 74 | 0.891 | 10,712,102 | 78,866 |
| Denmark | 137 | 88 | 0.930 | 5,795,391 | 42,430 |
| Finland | 18 | 86 | 0.925 | 5,542,073 | 303,890 |
| city of Fortaleza, Ceará, Brazil | 7,786 | 100 | 0.754 | 2,686,612 | 312 |
| France | 119 | 82 | 0.891 | 65,296,176 | 547,557 |
| Germany | 240 | 76 | 0.939 | 83,825,861 | 348,560 |
| Greece | 81 | 85 | 0.870 | 10,414,904 | 128,900 |
| Hungary | 107 | 72 | 0.845 | 9,656,450 | 90,530 |
| Ireland | 72 | 63 | 0.942 | 4,946,213 | 68,890 |
| Italy | 206 | 69 | 0.883 | 60,447,728 | 294,140 |
| Japan | 347 | 92 | 0.915 | 126,414,795 | 364,555 |
| Kuwait | 240 | 100 | 0.808 | 4,280,111 | 17,820 |
| Macedonia | 83 | 59 | 0.759 | 2,083,360 | 25,220 |
| city of Manaus, Amazonas, Brazil | 158 | 100 | 0.737 | 2,219,580 | 11,401 |
| Netherlands | 508 | 92 | 0.934 | 17,140,821 | 33,720 |
| New York City | 10,194 | 100 | 0.941 | 8,336,817 | 784 |
| Norway | 15 | 83 | 0.954 | 5,427,784 | 365,268 |
| Portugal | 111 | 66 | 0.850 | 10,191,976 | 91,590 |
| city of Rio de Janeiro, RJ, Brazil | 5,266 | 100 | 0.799 | 6,747,815 | 1,200 |
| Russia | 9 | 74 | 0.824 | 145,944,331 | 16,376,870 |
| Slovenia | 103 | 55 | 0.902 | 2,078,983 | 20,140 |
| South Korea | 527 | 82 | 0.906 | 51,276,136 | 97,230 |
| Spain | 94 | 80 | 0.893 | 46,757,635 | 498,800 |
| State of Acre | 4 | 73 | 0.663 | 894,470 | 164,124 |
| State of Amazonas | 2 | 79 | 0.674 | 4,207,714 | 1,559,169 |
| State of Pará | 6 | 68 | 0.646 | 8,602,865 | 1,245,871 |
| State of Roraima | 2 | 76 | 0.707 | 631,181 | 223,645 |
| Sweden | 25 | 88 | 0.937 | 10,109,031 | 410,340 |
| Switzerland | 219 | 74 | 0.946 | 8,664,406 | 39,516 |
| Tokyo, Japan | 6,158 | 100 | 0.941 | 13,491,000 | 2,191 |
| United Kingdom | 279 | 83 | 0.920 | 67,886,011 | 241,930 |
| **Not Controlled areas** |  |  |  |  |  |
| Argentina | 17 | 93 | 0.83 | 45,259,525 | 2,736,690 |
| Australia | 3 | 86 | 0.938 | 25,545,026 | 7,682,300 |
| Belarus | 47 | 79 | 0.817 | 9,448,832 | 202,910 |
| city of Belo Horizonte, MG, Brazil | 7167 | 100 | 0.81 | 2,521,564 | 331 |
| Bosnia and Herzegovina | 64 | 52 | 0.769 | 3,277,541 | 51,000 |
| Bulgaria | 64 | 76 | 0.861 | 6,940,012 | 108,560 |
| Chile | 26 | 85 | 0.847 | 19,141,470 | 743,532 |
| Colombia | 46 | 80 | 0.761 | 50,965,881 | 1,109,500 |
| Costa Rica | 100 | 80 | 0.794 | 5,101,269 | 51,060 |
| Croatia | 73 | 58 | 0.837 | 4,101,200 | 55,960 |
| Brasília, FD Brazil | 444.66 | 96.62 | 0.824 | 3,055,149 | 5,761 |
| Israel | 400 | 93 | 0.906 | 9,197,590 | 21,640 |
| Lebanon | 667 | 78 | 0.73 | 6,820,558 | 10,230 |
| Libya | 4 | 78 | 0.708 | 6,885,460 | 1,759,540 |
| Luxembourg | 242 | 88 | 0.909 | 627,509 | 2,590 |
| Moldova | 105 | 43 | 0.711 | 4,032,473 | 32,850 |
| Oman | 16 | 87 | 0.834 | 5,125,566 | 309,500 |
| Peru | 26 | 79 | 0.759 | 33,041,424 | 1,280,000 |
| Poland | 124 | 60 | 0.872 | 37,840,045 | 306,230 |
| city of Porto Alegre, RS, Brazil | 2837.53 | 100 | 0.805 | 1,488,252 | 495 |
| Qatar | 248 | 96 | 0.848 | 2,807,805 | 11,610 |
| Romania | 84 | 55 | 0.816 | 19,217,049 | 230,170 |
| city of São Paulo, SP, Brazil | 7398.26 | 100 | 0.805 | 12,325,232 | 1,521 |
| Saudi Arabia | 16 | 84 | 0.857 | 34,895,566 | 2,149,690 |
| Serbia | 100 | 56 | 0.799 | 8,731,751 | 87,460 |
| state of Alagoas | 112.23 | 73.64 | 0.631 | 3,351,543 | 27,843 |
| state of Amapá | 4.69 | 89.81 | 0.708 | 861,773 | 142,471 |
| state of Bahia | 24.82 | 72.07 | 0.66 | 14,930,634 | 564,760 |
| state of Ceará | 56.76 | 75.09 | 0.682 | 9,187,103 | 148,894 |
| state of Espírito Santo | 76.25 | 85.29 | 0.74 | 4,064,052 | 46,074 |
| state of Goiás | 17.65 | 90.29 | 0.735 | 7,113,540 | 340,203 |
| state of Maranhão | 19.81 | 63.07 | 0.639 | 7,114,598 | 329,642 |
| state of Mato Grosso | 3.36 | 81.9 | 0.725 | 3,526,220 | 903,207 |
| state of Mato Grosso do Sul | 6.86 | 85.64 | 0.729 | 2,809,394 | 357,146 |
| state of Minas Gerais | 33.41 | 83.38 | 0.731 | 21,292,666 | 586,521 |
| state of Paraíba | 66.7 | 75.37 | 0.658 | 4,039,277 | 56,467 |
| state of Paraná | 52.4 | 85.31 | 0.749 | 11,516,840 | 199,299 |
| state of Pernambuco | 89.63 | 80.15 | 0.673 | 9,616,621 | 98,068 |
| state of Piauí | 12.4 | 65.77 | 0.646 | 3,281,480 | 251,757 |
| state of Rio de Janeiro | 365.23 | 96.71 | 0.761 | 17,264,943 | 43,750 |
| state of Rio Grande do Norte | 59.99 | 77.82 | 0.684 | 3,534,165 | 52,810 |
| state of Rio Grande do Sul | 37.96 | 85.1 | 0.746 | 11,422,973 | 281,707 |
| state of Rondônia | 6.58 | 73.22 | 0.69 | 1,796,460 | 237,765 |
| state of Santa Catarina | 65.27 | 83.99 | 0.774 | 7,252,502 | 95,731 |
| state of São Paulo | 166.23 | 95.88 | 0.783 | 46,289,333 | 248,219 |
| state of Sergipe | 94.35 | 73.51 | 0.665 | 2,318,822 | 21,925 |
| state of Tocantins | 4.98 | 78.81 | 0.699 | 1,590,248 | 277,467 |
| Turkey | 110 | 76 | 0.807 | 84,477,895 | 769,630 |
| Ukraine | 75 | 69 | 0.75 | 43,691,576 | 579,320 |
| United Arab Emirates | 118 | 86 | 0.866 | 9,908,607 | 83,600 |
| United States of America | 36 | 82 | 0.92 | 331,303,997 | 9,834,000 |

### Comparisons

The restrictive analysis between controlled and not controlled areas yielded 33 appropriate comparisons, as shown in Table 2. Only one comparison out of 33 (3%) - state of Roraima (Brazil) versus state of Rondonia (Brazil) - was significant (p-value = 0.04). After correction for residual analysis, it did not pass the autocorrelation test (Lagrange Multiplier test=0.04). (Further details are in <u>Supplemental Material - Restrictive Analysis</u>).

**Table 2.** Comparisons using the 4-point criteria. Comparability was considered if at least 3 out of 4 of the following conditions were similar: a) population density, b) percentage of the urban population, c) Human Development Index and d) total area of the region. Similarity was considered adequate when a variation in conditions a), b) and c) was within 30%, while, for condition d), a variation of 50% was considered adequate (Further details are in <u>Data sharing - 4 point criteria</u>).

| Region/Country (controlled) | Region/Country (not controlled) | Comparability<sup>a</sup> | p-value<sup>b</sup> |
| --- | --- | --- | --- |
| Austria | Serbia | 4 | 0.055 |
| Bahrain | city of Porto Alegre, RS, Brazil | 4 | 0.911 |
| Belgium | Israel | 4 | 0.114 |
| Canada | Australia | 4 | 0.965 |
| Czech Republic | State of Alagoas | 3 | 0.3501 |
| Denmark | Turkey | 3 | 0.911 |
| Finland | State of Goiás | 4 | 0.268 |
| city of Fortaleza, CE, Brazil | city of Belo Horizonte, MG, Brazil | 4 | 0.301 |
| France | Ukraine | 3 | 0.623 |
| Germany | Qatar | 3 | 0.892 |
| Greece | Bulgaria | 4 | 0.275 |
| Ireland | Croatia | 4 | 0.711 |
| Italy | State of São Paulo | 3 | 0.928 |
| Japan | Israel | 3 | 0.102 |
| Kuwait | Luxembourg | 3 | 0.060 |
| city of Manaus, AM, Brazil | Qatar | 3 | 0.524 |
| Macedonia | Romania | 3 | 0.6169 |
| Netherlands | city of Brasília, Brazil | 3 | 0.459 |
| New York City | city of São Paulo, SP, Brazil | 3 | 0.645 |
| Norway | Saudi Arabia | 3 | 0.379 |
| Portugal | United Arab Emirates | 3 | 0.248 |
| Russia | United States of America | 3 | 0.557 |
| Slovenia | Poland | 4 | 0.875 |
| South Korea | Lebanon | 3 | 0.645 |
| Spain | State of Minas Gerais | 3 | 0.853 |
| State of Acre | State of Amapá | 4 | 0.803 |
| State of Amazonas | Colombia | 3 | 0.638 |
| State of Pará | Libya | 3 | 0.681 |
| State of Roraima | State of Rondônia | 3 | 0.042 |
| Sweden | State of Bahia | 4 | 0.131 |
| Switzerland | State of Espírito Santo | 3 | 0.745 |
| Tokyo, Japan | city of São Paulo, SP, Brazil | 4 | 0.731 |
| United Kingdom | State of Rio Grande do Sul | 3 | 0.084 |

<sup>a</sup> From four-point criteria, how many criteria were present
<sup>b</sup> Linear regression

The global comparison yielded 3,741 combinations; from these, 184 (4.9%) had a p-value < 0.05, after correcting for False Discovery Rate (Table S3). After performing the residual analysis, by testing for cointegration between response and covariate, normality of the residuals, presence of residual autocorrelation, homoscedasticity, and functional specification, only 63 (1.6%) of models passed all tests (Table S4). Closer inspection of several cases where the model did not pass all the tests revealed a common factor: the presence of outliers, mostly due to differences in the epidemiological week in which deaths started to be reported. A heat map showing the comparison between the 87 regions is presented in Figure 2.

**Figure 2.**
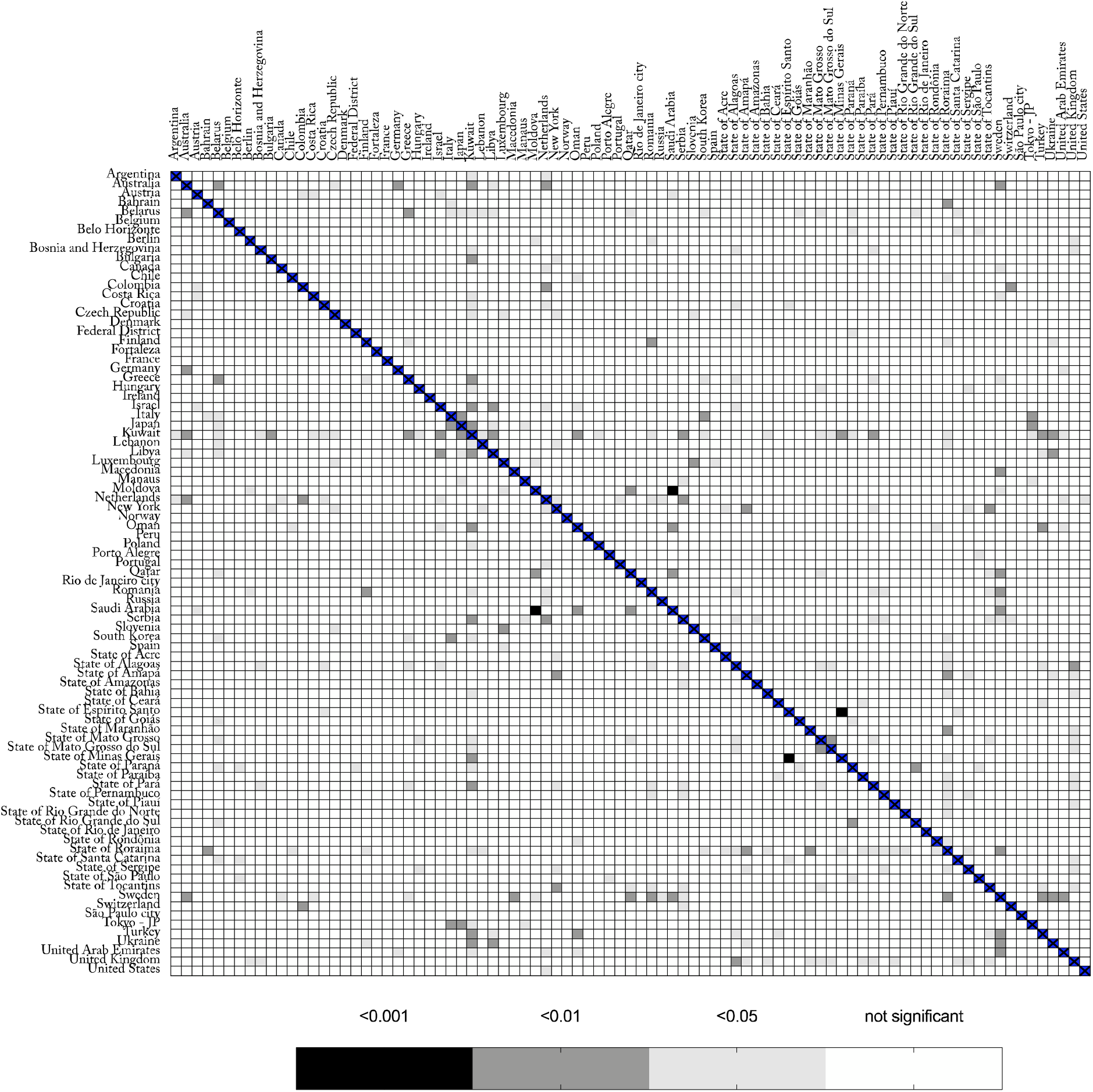
Heat map comparing different regions with COVID-19. The bar below represents p-values for the linear regression.

## Discussion

We were not able to explain the variation of deaths/million in different regions in the world by social isolation, herein analyzed as differences in staying at home, compared to baseline. In the restrictive and global comparisons, only 3% and 1.6% of the comparisons were significantly different, respectively. These findings are in accordance with those found by Klein et al. (Klein et al.). These authors explain why lockdown was the least probable cause for Sweden’s high death rate from COVID-19 (Klein et al.). Likewise, Chaudry et al. made a country-level exploratory analysis, using a variety of socioeconomic and health-related characteristics, similar to what we have done here, and reported that full lockdowns and wide-spread testing were not associated with COVID-19 mortality per million people (Chaudhry et al. 2020). Different from Chaudry et al., in our dataset, after 25 epidemiological weeks, (counting from the 9^th^ epidemiological week onwards in 2020) we included regions and countries with a “plateau” and a downslope phase in their epidemiological curves. Our findings are in accordance with the dataset of daily confirmed COVID-19 deaths/million in the UK. Pubs, restaurants, and barbershops were open in Ireland on June 29^th^ and masks were not mandatory (Therese 2020); after more than 2 months, no spike was observed; indeed, death rates kept falling (Daily confirmed COVID-19 deaths per million, rolling 7-day average). Peru has been considered to be the most strict lockdown country in the world (Tegel 2020), nevertheless, by September 20^th^, it had the highest number of deaths/million (Coronavirus Update (Live): 31,036,957 Cases and 962,339 Deaths from COVID-19 Virus Pandemic - Worldometer). Of note, differences were also observed between regions that were considered to be COVID-19 controlled, e.g., Sweden versus Macedonia. Possible explanations for these significant differences may be related to the magnitude of deaths in these countries.

Our results are different from those published by Flaxman et al. These authors calculated that NPIs would prevent 3.1 million deaths across 11 European countries (Flaxman et al. 2020). The discrepant results can be explained by different approaches to the data. While Flaxman et al. assumed a constant reproduction number (R_t_) to calculate the total number of deaths, which eventually did not occur, we calculated the difference between the actual number of deaths between 2 countries/regions. The same explanation for the discrepancy can be applied to other publications where mathematical models were created to predict outcomes (Ambikapathy and Krishnamurthy 2020; Ibarra-Vega 2020; Liu et al. 2020; Nussbaumer-Streit et al. 2020; Sjödin et al. 2020). Most of these studies dealt with COVID-19 cases (Banerjee and Nayak 2020; Wang et al. 2020) and not observed deaths. Despite its limitations, reported deaths are likely to be more reliable than new case data. Further explanations for different results in the literature, besides methodological aspects, could be explained by the complexity of the virus dynamic and its interaction with the environment. It is unwise to try to explain a complex and multifactorial condition, with the inherent constant changes, using a single variable. An initial approach would employ a linear regression to verify the influence of one factor over an outcome. Herein we were not able to identify this association.

This study has limitations. Different from the established paradigm of randomized clinical trial, this is an ecological study. An ecological study observes findings at the population level and generates hypotheses (Pearce 2000). Population-level studies play an essential part in defining the most important public health problems to be tackled (Pearce 2000), which is the case here. Another limitation was the use of Google Community Mobility Reports as a surrogate marker for staying at home. This may underestimate the real value: for instance, if a user’s cell phone is switched off while at home, the observation will be absent from the database. Furthermore, the sample does not represent 100% of the population. This tool, nevertheless, has been used by other authors to demonstrate the efficacy in reducing the number of new cases after NPI (Delen et al. 2020; Vokó and Pitter 2020). Using different methodologies for measuring mobility may introduce bias and would prevent comparisons between different countries. The number of deaths may be another issue. Death figures may be underestimated, however, reported deaths may be more relevant than new case data. The arbitrary criteria used for including countries and regions, the restrictive comparisons, and our definition of an area as COVID-19 controlled are open for criticism. Nonetheless, these arbitrary criteria were created a priori to the selection of the countries. With these criteria, we expected to obtain representative regions of the world, compare similar regions, and obtain accurate data. By using a HAQI of ≥ 67, we assumed that data from these countries would be accurate, reliable, and health conditions were generally good. Nevertheless, the global analysis of the regions (*n* = 3741 comparisons) overcame any issue of the restrictive comparison. Indeed, the global comparison confirmed the results found in the restrictive one; only 1.6% of the death rates could be explained by staying at home. Also, our effective sample size in all studies is only 25 epidemiological weeks, which is a very small sample size for a time series regression. The small sample size and the non-stationary nature of COVID-19 data are challenges for statistical models, but our analysis, with 25 epidemiological weeks, is relatively larger than previous publications which used only 7 weeks (Ghosal et al. 2020). The effects of small samples in this case are related to possible large type II errors and also affect the consistency of the ordinary least square estimates. Nevertheless, given the importance of social isolation promoted by world authorities (COVID-19 advice - Physical distancing), we expected a higher incidence of significant comparisons, even though it could be an ecological fallacy. The low number of significant associations between regions for mortality rate and the percentage of staying at home may be a case of exception fallacy, which is a generalization of individual characteristics applied at the group-level characteristics (Miller and Brewer 2003).

There are strengths to highlight. Inclusion criteria and the Healthcare Access and Quality Index were incorporated. We obtained representative regions throughout the world, including major cities from 4 different continents. Special attention was given to compiling and analyzing the dataset. We also devised a tailored approach to deal with challenges presented by the data. To our knowledge, our modeling approach is unique in pooling information from multiple countries all at once using up-to-date data. Some criteria, such as population density, percentage of urban population, HDI, and HAQI, were established to compare similar regions. Finally, we gave special attention to the residual analysis in the linear regression, an absolutely essential aspect of studies using small samples.

In conclusion, using this methodology and current data, in ∼98% of the comparisons using 87 different regions of the world we found no evidence that the number of deaths/million is reduced by staying at home. Regional differences in treatment methods and the natural course of the virus may also be major factors in this pandemic, and further studies are necessary to better understand it.

## Supporting information

checklist

## Data Availability

Supplemental Material:
The Python and R scripts, raw data, supplemental Tables are available.

https://drive.google.com/drive/folders/1239llmxz9YenWweWXA1wgdf07WFYDrYG

https://gist.github.com/rsavaris66/eccfc6caf4c9578d676c134fac74d3fe

## Supplemental Material

The Python and R scripts are available at https://gist.github.com/rsavaris66/eccfc6caf4c9578d676c134fac74d3fe

More supplemental material, including raw data, is available at this https://drive.google.com/drive/folders/1239llmxz9YenWweWXA1wgdf07WFYDrYG

## SUPPLEMENTAL MATERIAL

### Approach for analyzing the time series data

**Figure S1.**
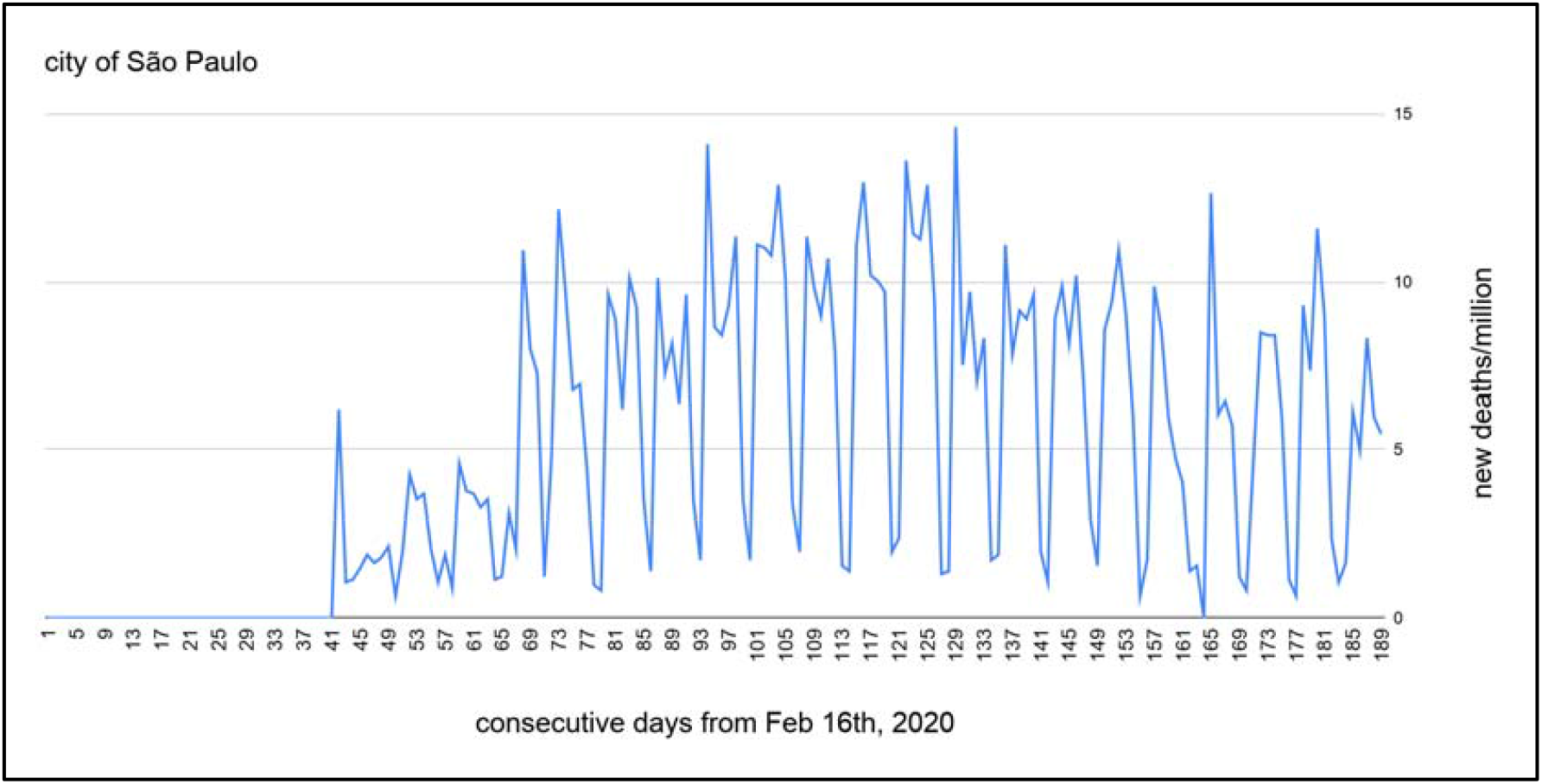
Characteristics of the time series data on new daily deaths/million in the city of São Paulo over 187 days. Note the non-stationary time series pattern.

**Figure S2.**
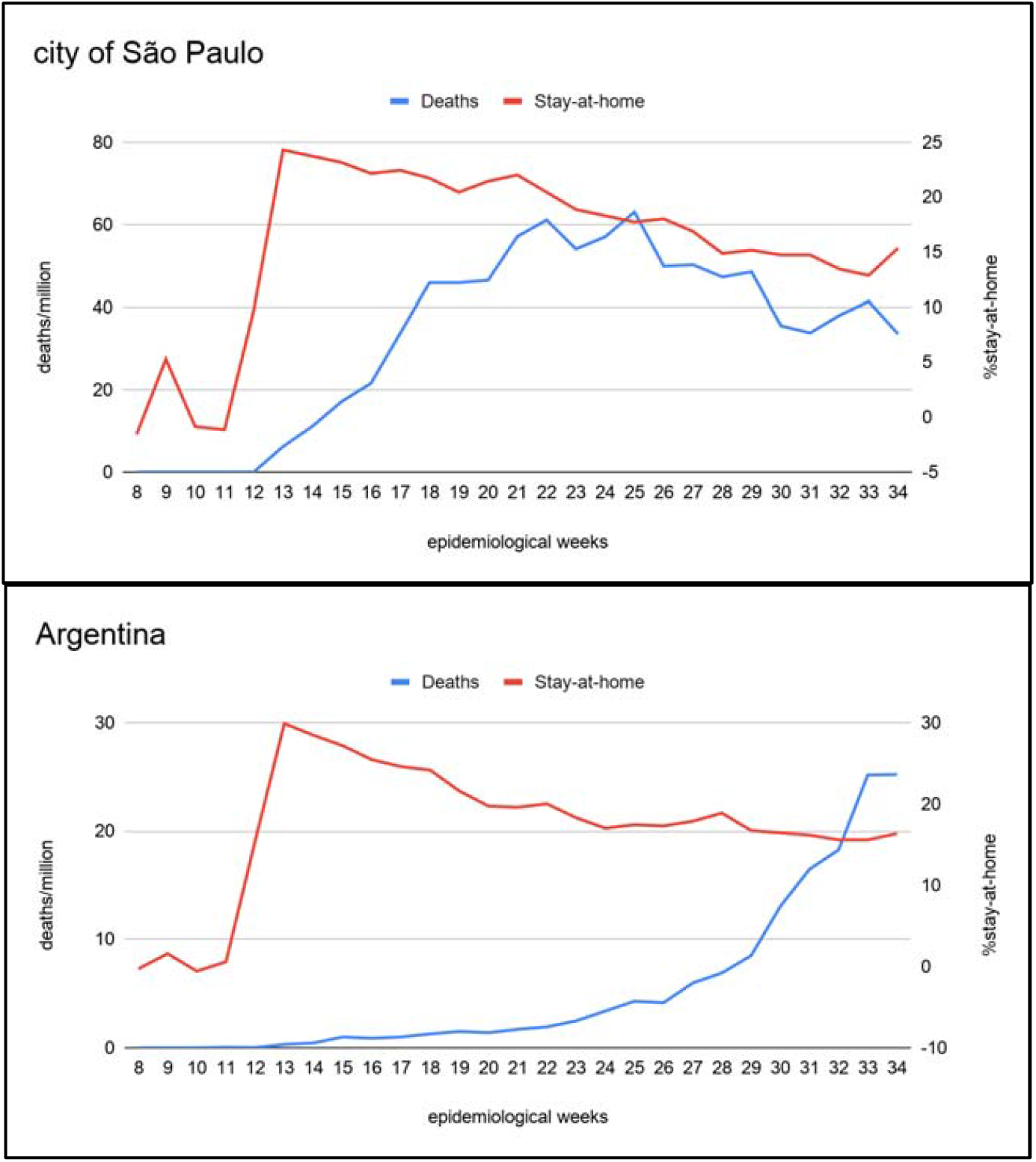
Data aggregation of the number of deaths/million in the city of São Paulo and in Argentina over several epidemiological weeks, compared to the percentage of staying at home. Data aggregation by epidemiological week is a plausible alternative. In this way, artificial seasonality, imposed by work scheduled during weekends and the effect of governmental control over social interaction, in a regression framework, are mitigated. The drawback is that the sample size is significantly reduced from 187 days (Figure S1) to 26 epidemiological weeks.

### Definition of areas with and without controlled cases of COVID-19

Regions were classified as controlled for cases of COVID-19 if they present at least 2 out of the 3 following conditions: a) type of transmission classified as “clusters of cases”, b) a downward curve of newly reported deaths in the last 7 days, and c) a flat curve in the cumulative total number of deaths in the last 7 days (variation of 5%) according to the World Health Organization (WHO Coronavirus Disease (COVID-19) Dashboard). An example is shown in Figure S3.

**Figure S3.**
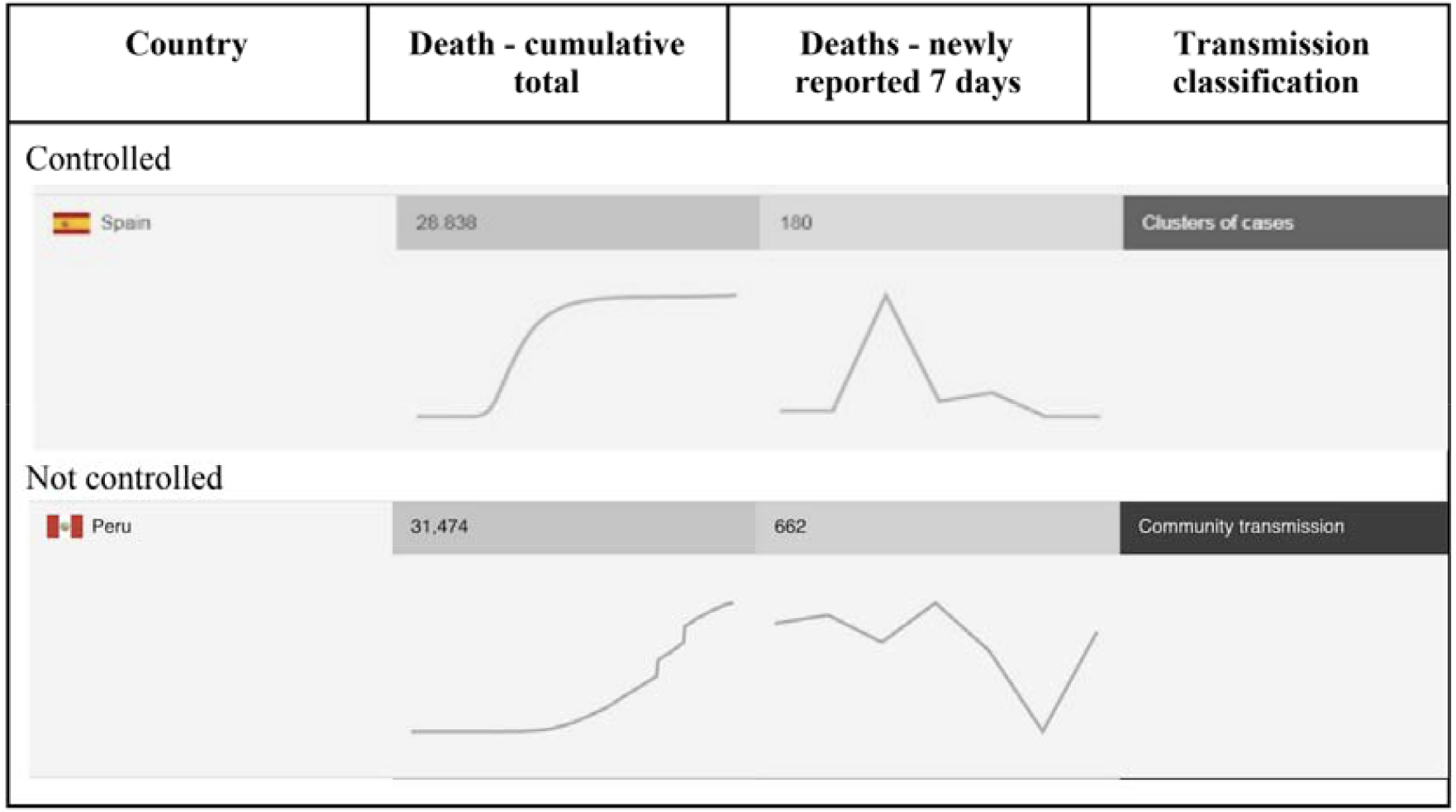
Example of areas with and without control of COVID-19

<u>Table S3).</u> Comparison between 184 countries and regions displaying a significant association between the variation of number of deaths/million and the variation of the percentage of staying at home, after False Discovery Rate (FDR B-H) analysis. Intercept and coef_isolation are β0 and β1 from the linear regression formula; Shapiro is the test for normality; White is the test for heteroskedasticity; LM is the Lagrange Multiplier test for autocorrelation, Reset is the Ramsey’s RESET test for functional specification (all should have a p-value ≥0.05 for a valid comparison), Coint= the p-value of the Phillips-Perron test applied to the residual of the regression (for a valid comparison, p-value<0.05)

<u>Table S4)</u> Comparison between 63 countries and regions displaying a significant association between the variation of number of deaths/million and the variation of the percentage of staying at home, after False Discovery Rate analysis and after residual analysis. Intercept and coef_isolation are β0 and β1 from the linear regression formula; Shapiro is the test for normality; White is the test for heteroskedasticity; LM is the Lagrange Multiplier test for autocorrelation, Reset is the Ramsey’s RESET test for functional specification (all should have a p-value ≥0.05 for a valid comparison), Coint= the p-value of the Phillips-Perron test applied to the residual of the regression (for a valid comparison, p-value<0.05).

